# Associations between area-level health-related social factor indices and risk of acute COVID-19: An EHR-based cohort study from the RECOVER program

**DOI:** 10.1101/2022.12.02.22282944

**Authors:** Deena J. Chisolm, Ryan Webb, Katherine S. Salamon, Julia Schuchard, Eneida A Mendonca, Marion R. Sills, Payal B Patel, Jordan Musante, Christopher B. Forrest, Ravi Jhaveri, Nathan M Pajor, Suchitra Rao, Grace M. Lee, Asuncion Mejias

## Abstract

**Background:** Research demonstrates that SARS-CoV-2 infection (COVID-19) among adults disproportionately impacts racial and ethnic minorities and those living in lower-income communities. Similar research in children is limited due, in part, to the relatively low COVID-19 incidence in children compared to adults. This analysis, conducted as part of the RECOVER Initiative, explores this question.

**Methods:** Electronic health record (EHR) data from PEDSnet, a multi-institutional research network of pediatric healthcare organizations, were geocoded and linked to two indices of contextual social deprivation: the Area Deprivation Index and the Child Opportunity Index. Univariate statistics were employed to test the association between each index and COVID19 positivity among children ages 0-20 tested at one of six Children’s hospitals. Multivariate logistic regression was used to explore the relationship between these social context indices and racial disparities in positivity, controlling co-variates.

**Results:** Both ADI and COI were significantly associated with COVID-19 positivity in univariate and adjusted models, particularly in the pre-delta and delta variant waves. ADI showed a stronger association. Higher rates of positivity were found for non-Hispanic Black, Hispanic, and multi-racial children compared to non-Hispanic White children. These racial disparities remained significant after control for either index and for other variables.

**Conclusion:** ADI and COI are significantly associated with COVID-19 test positivity in a population of children and adolescents tested in children’s hospital settings. These social contextual variables do not fully explain racial disparities, arguing that racial disparities are not solely a reflection of socioeconomic status. Future disparities research should consider both race and social context.

## INTRODUCTION

Since the onset of the global pandemic of SARS-CoV-2 in 2020, much has been written regarding the influence of the social environment on risk and severity of acute SARS-CoV-2 infection (1-3). Consistently, such research demonstrates that SARS-CoV-2 infection (COVID-19) among adults disproportionately impacts racial and ethnic minorities, those living in lower-income communities, those working in jobs with increased exposure risk, and those with reduced access to healthcare. While some of the noted racial and ethnic disparities are driven by contextual health-related social factors, commonly referred to as social determinants of health (SDoH), independent race effects have also been identified (4-8).

Research in children is limited due, in part, to the fact that the incidence and severity of acute SARS-CoV-2 infection has been low in comparison with adults (9). Detailed analyses of the associations of social factors and race with pediatric COVID-19, severity, mortality, and long-term effects, requires large, diverse cohorts of children and adolescents that have not been available for research to date.

As part of the NIH Researching COVID to Enhance Recovery Initiative (RECOVER; https://recovercovid.org/), this paper uses a pediatric EHR cohort (PEDSnet) to 1) investigate the relationship among community-level SDoH and diagnosed COVID-19 in children and adolescents < 21 years of age; 2) determine the role of health-related social factors in explaining racial and ethnic disparities; and 3) explore the best approaches for future research on social factors associated with acute and post-acute COVID-19.

## METHODS

### Data Source

This analysis was conducted as part of the RECOVER Initiative. Established In 2021 by the National Institutes of Health (NIH), the RECOVER Initiative has the goal of harnessing research knowledge and infrastructure across the country to understand, treat, and prevent post-acute sequelae of COVID-19 (PASC), commonly referred to as “long COVID”. PASC is defined as ongoing, relapsing, or new symptoms, or other health effects occurring four or more weeks after an acute SARS-CoV-2 infection (10). The RECOVER consortium represents and supports more than 100 researchers who are leading studies on PASC at more than 200 institutions around the country (10-16). Study participants are socio-demographically and clinically diverse and include adults, pregnant people, and children. One component of this multifaceted consortium is PEDSnet, a multi-institutional research network that analyzes electronic health record (EHR) data from several of the nation’s largest pediatric healthcare organizations. The rich PEDSNet data offers sufficient size and diversity to explore associations between the social environments of children and the risk of COVID-19.

Participating clinical sites include six children’s hospitals that are members of PEDSnet and have census block group data available for geocoding: Children’s Hospital of Philadelphia (CHOP), Cincinnati Children’s Hospital Medical Center, Children’s Hospital Colorado, Nationwide Children’s Hospital in Columbus, Nemours Children’s Health System (a Delaware and Florida health system), Seattle Children’s Hospital, and Stanford Children’s Health. PEDSnet data is harmonized and stored using the PEDSnet common data model (17). Children’s Hospital of Philadelphia’s Institutional Review Board designated this study as not human subjects’ research and waived informed consent.

### Study Cohorts

This analysis included children and adolescents, ages 0-20, who underwent COVID-19 testing within the PEDSnet network between March 2020 and September 2022. Children were classified into positive and negative cohorts. The positive cohort included youths with an incident positive SARS-CoV-2 viral test (antigen, polymerase chain reaction [PCR], or serology reflecting past SARS-CoV-2 infection) (15, 18). The negative cohort was defined as patients with only negative test results during the study period.

### Primary Outcome Variable

The primary outcome variable is COVID-19 positivity (yes/no).

### Primary Independent Variables

#### Health-related social factors

Geocoded PEDSnet EHR data were linked to data from two indices measuring neighborhood level socio-economic and environmental resources and exposures, the Area Deprivation Index (ADI) and the Child Opportunity Index (COI) (19, 20), at the Census block group level. The ADI has been refined from a measure created by the Health Resources and Services Administration over 30 years ago. It includes the domains of income, education, employment, and housing quality, and has been validated for neighborhood comparisons at the Census block group level. Predictive validity assessment documented correlations above 0.40 with health outcomes including all-cause mortality, infant mortality, and low birth weight. The COI 2.0, launched in 2020, is an index of neighborhood resources and conditions that support healthy development. It includes 29 indicators representing three domains: education, health and environment, and social and economic. Assessment of predictive validity found that COI 2.0 predicted 43% of the variance in life expectancy at the neighborhood level and at least half of the variance in adult outcomes measured by the 500 Cities Data and the Opportunity Index. The COI does not report data for census tracts missing more than 50% of the indicators in any of the three domains, so some census tracts included in this analysis do not map to a COI.

Census block group was derived from a patient’s current primary address only, using 2020 census boundaries, and the most recent derivations for each of ADI (2020) and COI (2015) were used in this analysis. To link COI data, 2020 census block groups were linked to 2010 census tracts where the 2020 block group is contained entirely within a 2010 tract (19). ADI can be mapped to all US census blocks; however, COI is limited to Census blocks in the 500 largest U.S cities. There is substantial overlap of data sources and indicators between the two indices, yet they differ in the population of interest (general vs. pediatric), data sources, and weighting mechanisms. Higher scores on the ADI reflect higher levels of deprivation, while higher scores on the COI reflect higher social opportunity. As such, the scales appear reversed in association analyses. COI has lower coverage of census block groups and will only match for a subset of the cohort. For both indices, scores were grouped into quintiles using national normative values.

#### Race/Ethnicity

Race and ethnicity were determined using data available in each patient’s EHR. Children are classified as Non-Hispanic White, Non-Hispanic Black, Non-Hispanic Asian American/Pacific Islander(AAPI), multiple races, or other group/not available. Specific methods for collection of this data may vary across PEDSnet institutions. For this analysis, race and ethnicity are not hypothesized as biologic variables but rather as markers of historical and current inequities that impact minoritized populations.

#### Pandemic Wave

The pandemic period was classified into three waves. Pre-Delta (March 2020 through June 2021), Delta (July 2021 through December 2021), and Omicron (January 2022 through September 2022). We recognize that these waves are not fully distinct but instead represent the predominant variants in each period. Entrance data is used to determine a single wave for each patient. In the positive cohort, entrance date is the date of their first positive test. For patients in the negative group with more than one negative test, one negative test was randomly selected to define the cohort entrance date.

### Co-variates

Multivariable models adjust for potential confounding from covariates including gender (male, female, other/unknown), age at cohort entrance date, and presence of chronic health conditions. The Pediatric Medical Complexity Algorithm (PMCA) Version 2.0 was applied to categorize children as having no chronic conditions, non-complex chronic conditions, or complex chronic comorbidities, considering diagnoses up to 3 years before cohort entrance. Neither data for individual nor community level vaccination status were available for this analysis.

### Analyses

We first provide descriptive frequencies for the study population and calculate the percent positive overall and by sociodemographic and clinical characteristics. We tested differences in positivity for each characteristic using chi-square tests and calculated effect sizes using the standardized differences. The latter may be interpreted in terms of standard deviations (e.g., 0.5 = one-half the standard deviation of the variable). Small, medium, and large effect sizes roughly correspond to 0.2, 0.5, and 0.8, respectively. We used logistic regression models with COVID-19 test positivity (yes/no) as the outcome to examine associations with the ADI or COI and the following covariates: race/ethnicity, age, gender, pandemic wave, and presence of chronic health conditions (21, 22). We also tested interaction effects between race and each index to examine associations among race, ADI/COI, and COVID-19 test positivity.

## RESULTS

Our analysis included 836,750 unique patients tested for COVID-19 and successfully matched to an ADI score via geocoding. Forty-seven percent of patients were female. The racial distribution was 49.3% non-Hispanic White, 14.5% African American or Black, 17.2% Hispanic, 4.7% Asian American/Pacific Islander, and 3.7% multiracial. Race was classified as other or unknown for 10.9 percent of youth. Nearly half of the study population (45.8%) was tested in the “pre-Delta” wave, while 28.2% were tested during the Delta wave and 26.0% were tested during the Omicron wave (**Table 1**). In total, 136,624 (16.3%) had at least one positive COVID-19 antigen or PCR test. Positivity rates were highest in the Omicron pandemic phase compared to the earlier periods. The positive and negative groups were statistically significantly different for each characteristic in Table 1 (p <.01), which is expected due to the large sample sizes. Positivity was significantly higher for non-Hispanic Black youth (19.9%) and Hispanic youth (18.6%), compared to white youth (15.0%). Age and sex differences were negligible. Tests conducted in the ED had the highest positivity rate (21.6%) and some variance was seen by hospital, with rates ranging from 12.9% to 19.2%.

**Table 1:** Demographic Characteristics of the Study Cohorts.

| **Characteristic** | **Negative** | **Positive** | **% Positive** | **Standardized difference** |
| --- | --- | --- | --- | --- |
|  | n (%) | n (%) |  |  |
| Total | 700108 | 136642 |  |  |
| **Age (years)** |  |  |  | 0.06 |
| 0-4 | 275299<br>(39.3) | 53955<br>(39.5) | 16.4% |  |
| 12-15 | 107641<br>(15.4) | 22276<br>(16.3) | 17.1% |  |
| 16-20 | 85808<br>(12.3) | 18631<br>(13.6) | 17.8% |  |
| 5-11 | 231360<br>(33.0) | 41780<br>(30.6) | 15.3% |  |
| **Sex** |  |  |  | 0.02 |
| Female | 331927<br>(47.4) | 65837<br>(48.2) | 16.6% |  |
| Male | 368035<br>(52.6) | 70794<br>(51.8) | 16.1% |  |
| Other/Unknown | 146<br>(<0.1) | 11<br>(<0.1) | 7.0% |  |
| **Language** |  |  |  | 0.03 |
| English language | 639447<br>(91.3) | 123896<br>(90.7) | 16.2% |  |
| Other/Unknown | 24997<br>(3.6) | 4767<br>(3.5) | 16.0% |  |
| Spanish language | 35664<br>(5.1) | 7979<br>(5.8) | 18.3% |  |
| **Pandemic Period** |  |  |  | 0.43 |
| Delta | 199595<br>(28.5) | 36171<br>(26.5) | 15.3% |  |
| Omicron | 160768<br>(23.0) | 56688<br>(41.5) | 26.1% |  |
| Pre-Delta | 339745<br>(48.5) | 43783<br>(32.0) | 11.4% |  |
| **PMCA Complex/Chronic** |  |  |  | 0.01 |
| Chronic | 93966<br>(13.4) | 17925<br>(13.1) | 16.0% |  |
| Complex | 79256<br>(11.3) | 16034<br>(11.7) | 16.8% |  |
| None | 526886<br>(75.3) | 102683<br>(75.1) | 16.3% |  |
| **Race/Ethnicity** |  |  |  | 0.15 |
| AAPI | 33017<br>(4.7) | 5473<br>(4.0) | 14.2% |  |
| Black or African American | 97506<br>(13.9) | 24224<br>(17.7) | 19.9% |  |
| Hispanic | 117438<br>(16.8) | 26831<br>(19.6) | 18.6% |  |
| Multiple race | 23973<br>(3.4) | 4687<br>(3.4) | 16.4% |  |
| Native American/Alaska Native | 1478<br>(0.2) | 259<br>(0.2) | 14.9% |  |
| Other/Unknown | 76309<br>(10.9) | 13399<br>(9.8) | 14.9% |  |
| White | 350387<br>(50.0) | 61769<br>(45.2) | 15.0% |  |
| **Test Location** |  |  |  | 0.21 |
| ED | 149527<br>(21.4) | 41091<br>(30.1) | 21.6% |  |
| Inpatient | 61215<br>(8.7) | 8682<br>(6.4) | 12.4% |  |
| Other/Unknown | 5345<br>(0.8) | 1171<br>(0.9) | 18.0% |  |
| Outpatient | 484021<br>(69.1) | 85698<br>(62.7) | 15.0% |  |

The study cohort distribution trended toward lower levels of deprivation than the general U.S. population, with 30.2% of patients in the lowest (least deprived) quintile and 10.9% in the highest (most deprived) quintile. If the cohort population were consistent with the general population, we would expect children to fall evenly into each quintile (20% each) given the use of national norms (Table 2).

**Table 2:** Social Indices by COVID Positivity: Area Deprivation Index and Child Opportunity Index.

| **Characteristic** | **Negative** | **Positive** | **% Positive** | **Standardized difference** |
| --- | --- | --- | --- | --- |
|  | n (%) | n (%) |  |  |
| **ADI** |  |  |  | 0.12 |
| Total | 700,108 | 136,642 |  |  |
| 0-20 | 216,879 (31.0) | 35,480 (26.0) | 14.1% |  |
| 21-40 | 191,162 (27.3) | 37,756 (27.6) | 16.5% |  |
| 41-60 | 132,505 (18.9) | 27,917 (20.4) | 17.4% |  |
| 61-80 | 85,393 (12.2) | 18,404 (13.5) | 17.7% |  |
| 81-100 | 74,169 (10.6) | 17,085 (12.5) | 18.7% |  |
| **COI** |  |  |  | 0.11 |
| Total | 559,434 | 109,937 |  |  |
| Very High | 201,209 (36.0) | 34,923 (31.8) | 14.8% |  |
| High | 109,520 (19.6) | 21,215 (19.3) | 16.2% |  |
| Moderate | 85,627 (15.3) | 17,093 (15.5) | 16.6% |  |
| Low | 72,908 (13.0) | 15,064 (13.7) | 17.1% |  |
| Very Low | 90,170 (16.1) | 21,642 (19.7) | 19.4% |  |

**Table 3:**
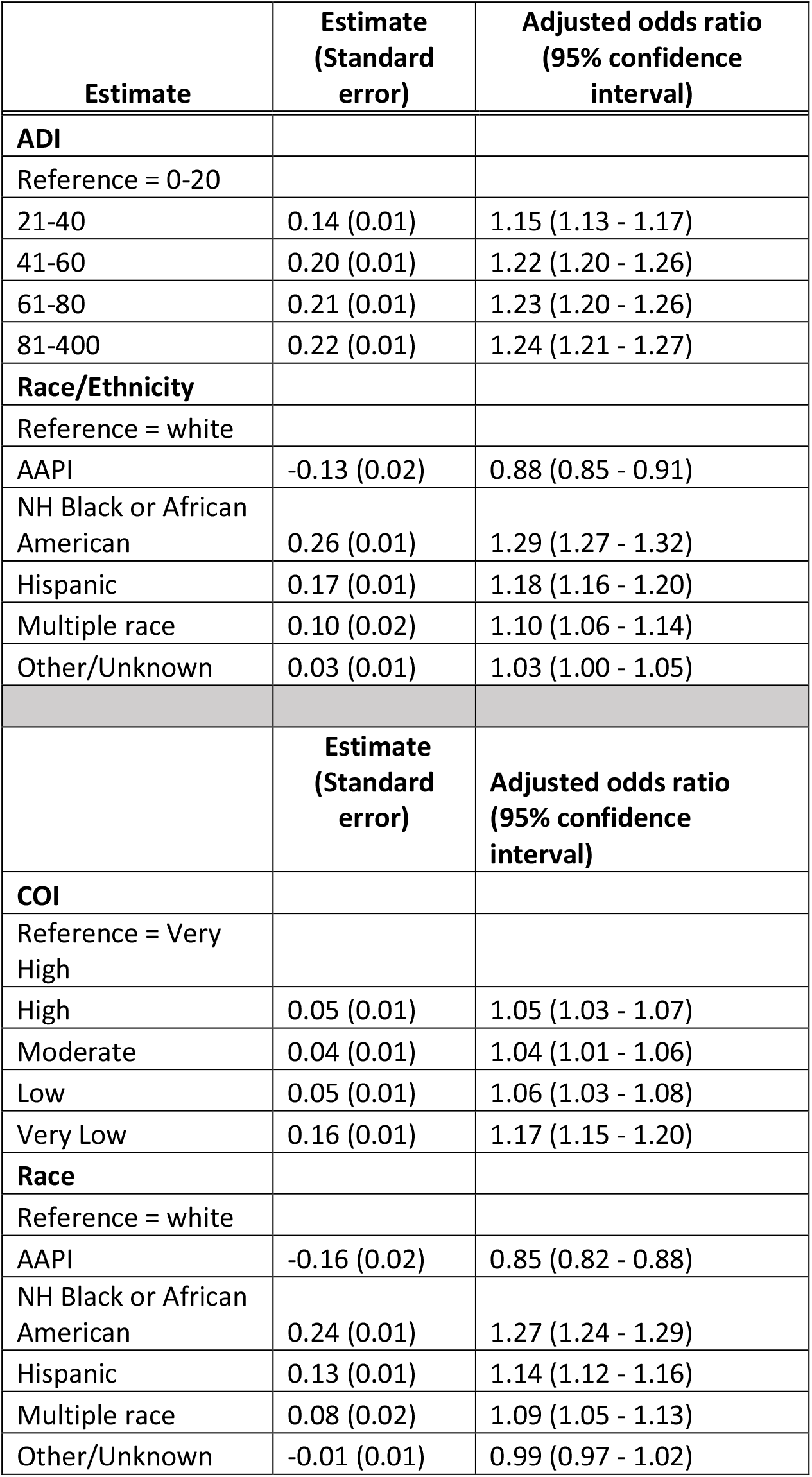
Logisitic Regression Models on Positivity by Race and SDH Controlling for Other Demographics.

Only 669,371 cohort members mapped to a COI (80.0%). Failure to match to COI was driven by the smaller number of indexed census blocks in the COI framework. The percentage of the cohort in the positive groups was no different than in the full (16.4%). In the sub-cohort matched to COI, however, only 19.5% of youth were in the quintile with the greatest opportunity (least deprivation).

Using either ADI or COI, we found that the rate of positivity increased as social disadvantage increased. For ADI, the positivity rate in the quintile of most deprivation was 24% higher than in the quartile of lowest deprivation (18.7% v 14.1%). Similarly in the COI, positivity was 30.1% higher in the quintile with the least social opportunity compared to the quintile with the most (19.4% vs. 14.8%). The relationship between positivity and disadvantage was stronger in the pre-Delta and Delta waves than in the Omicron wave (Figure 1).

**Figure 1.**
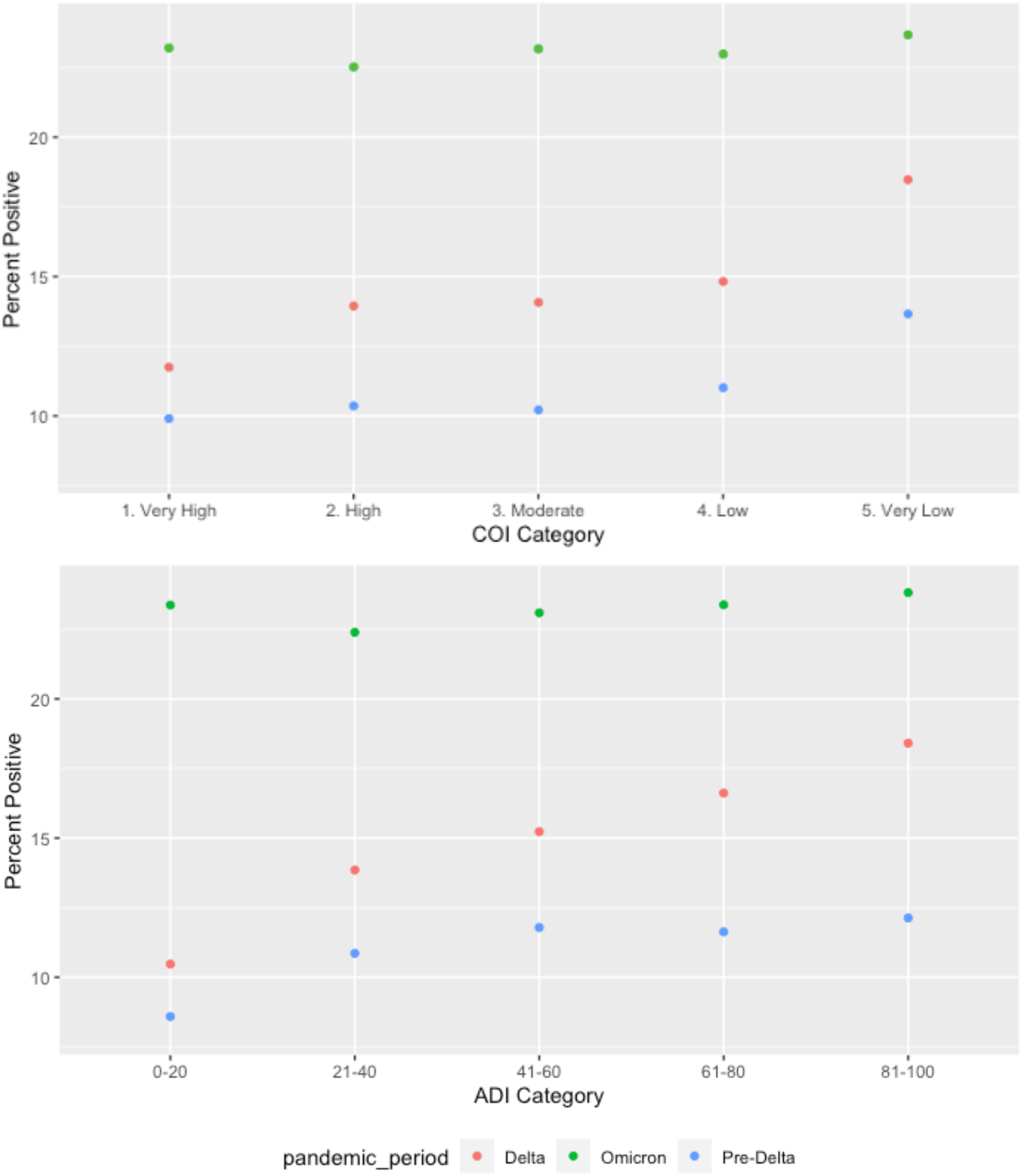
Positivity by disadvantage level in each pandemic wave

In multivariate analyses, SDoH remained significant in predicting positivity for both measures after controlling for co-variates. Logistic regression models found that the odds of positivity were higher in each quintile when compared to the least disadvantaged quintile. Neither ADI nor COI explained the racial and ethnic disparities found. The odds of positivity for non-Hispanic Black and Hispanic youth, compared to White, remained statistically significantly higher in the fully adjusted model. In the ADI model, odds ratios for non-Hispanic Black and Hispanic were 1.29 and 1.18, respectively. In the COI model, these odds ratios were 1.27 and 1.14.

Further analyses exploring the interactions between race and advantage show intriguing patterns (Figure 2). In the ADI model, while positivity increased with deprivation for the African American, Asian American/Pacific islander, and multiple race populations, the opposite was pattern was seen in Hispanic patients. White patients had positivity that increased as deprivation increased over the first three quintiles then descended. In the COI model, the associations between COI and COVID positivity for White and Hispanic children were more prominent compared to children in other racial/ethnic groups.

**Figure 2.**
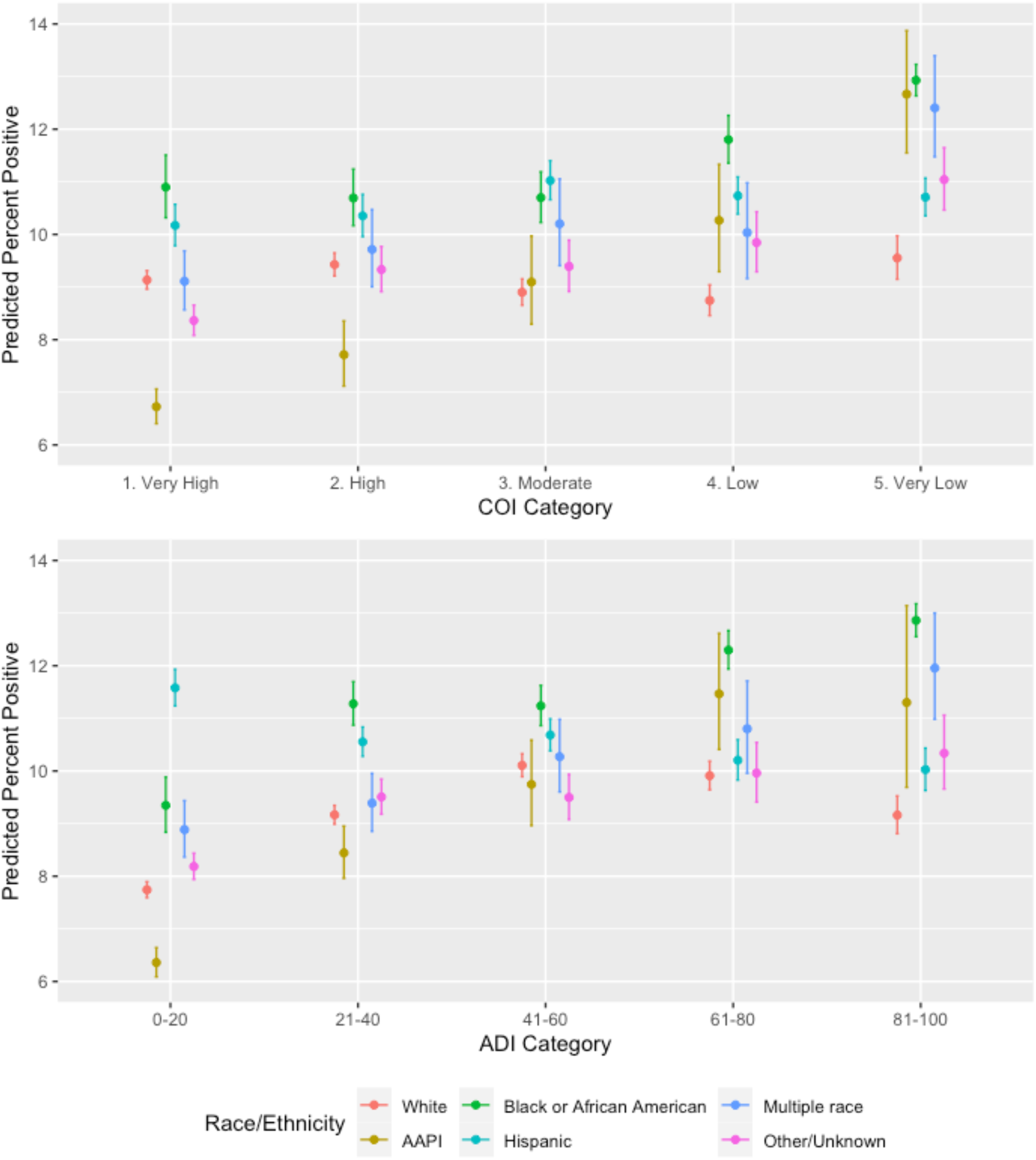
Race by ADI Interaction on Positivity

## DISCUSSION

There are structural inequities across the United States that have influenced how the COVID-19 pandemic has affected certain sub-populations. It is well established that social factors play a role in diagnosis, treatment, and outcomes of medical conditions. The COVID-19 pandemic has magnified health-related social factors impacting disparities in rates of COVID-19 and therefore, the development of PASC (23-25). Children bear the risk of being doubly impacted by their own health and their families’ health. This analysis highlights infection rates and predictors of infection, but also recognizes than an estimated 204,000 youth lost a caregiver due to COVID-19, with non-White children being more impacted. Black and Hispanic children lost a caregiver at nearly two times the rate of White children (26). Future studies would benefit from including information on child-level measures of stress, such as the adverse childhood events questionnaire, to factor in the impact of life-changing events, including those related to COVID-19, on clinical outcomes.

We found that both the ADI and the COI were significantly associated with the probability of a positive COVID-19 test in a population of children and adolescents tested in children’s hospital settings. Notably, the relationship between positivity and SDoH was concentrated in the pre-Delta and Delta phases of the pandemic. The Omicron phase, dominated by the most infectious variant seen to date, was much less driven by social factors in these analyses. During this phase of the pandemic, home testing practices were widely implemented, and Omicron infections were associated with less severe disease, likely leading to lower likelihood of care seeking at children’s hospitals, particularly for under-resourced families. As such, the weaker relationship between disadvantage indices and positivity in the Omicron wave could either suggest an actual weaker association or reflect a relative reduction in the number of children from disadvantaged communities tested at the RECOVER sties.

Comparing the two indices, there is a stronger association between COVID-19 and ADI. This is not a complete surprise given that the COI includes domains and variables, specific to children and youth, that are less directly linked to COVID-19 (e.g., third grade reading proficiency, college enrollment in nearby institutions) than the ADI’s primarily socio-economic factors. Differences may also be driven by the population characteristics of the census tracts excluded from the COI due to missing data.

Even with the significance of social factor indices, race/ethnicity remained a significant predictor, supporting the belief that racial inequities function in ways that are not fully captured by neighborhood measures of social disadvantage. This means that health-related social factors and race and ethnicity should both be studied to understand COVID19 disparities.

The link between neighborhood resources and infection mirrors results seen in adult populations. Consistent with our findings, Tung et al (27) found a significant association between COVID-19 positivity and ADI in a primarily adult population in Chicago in the pre-Delta phase. It can be argued that after most schools switched to virtual teaching to reduce the spread of COVID-19, adults in the household were the most likely source of infection. As a result, the caregivers’ risk factors became the child’s risk factors (28). This finding reminds us that while individual factors remain relevant, risk cannot be successfully addressed without attending to household and community behaviors and resources. This is particularly relevant for children who have little or no autonomy in their settings or exposures. Tung et al. also found that the ADI did not explain racial disparities in COVID-19 positivity rates. While there is no doubt that correlations between minoritized race membership and social deprivation exist and are rooted in current and historical interpersonal and systemic racism, other drivers of disproportional positivity include mistrust in public health messages, structural barriers to prevention modalities, and differential exposures. As such, race cannot be treated as a proxy for neighborhood social disadvantage nor can neighborhood social disadvantage be treated as a proxy for race. Both have a place in future research. The surprising interaction term for Hispanic ethnicity in our model further highlights the complexity of such relationships.

As future RECOVER studies explore associations between health-related social factors and PASC, this early work supports the importance of considering both neighborhood social disadvantage indices and race/ethnicity, the use of ADI as the generally preferred measure, and the importance of exploring interactions that may expose differential mechanisms across demographic sub-populations. Future research regarding health-related social factors should also include additional patient level variables, particularly a measure of childhood toxic stress to account for caregiver loss, though this work is not feasible within the existing EHR framework. The notable limitation of this analysis is the fact that the RECOVER pediatric EHR cohort is limited to children and adolescents who have received care at a RECOVER-participating site. These systems’ patient populations may not be representative of the nation. Children evaluated in these systems differ meaningfully from those seen in other types of healthcare settings and from those who did not access health care for similar conditions on measures including wealth, urbanicity, and insurance type. As such, our results likely underestimate the relationship between COVID-19 and health-related social factors and are not fully generalizable. We also note that ADI is measured at the block group level and does not necessarily reflect the social needs of individuals living in those geographies (ecological fallacy). Finally, we note that this analysis does not include adjustment for individual or community level vaccination status due to data availability. Even with these limitations, our findings document the strong relationship between acute COVID-19 and neighborhood social disadvantage and support the importance of targeted access to prevention, testing, vaccination, and treatment for pediatric populations in geographic areas with substantial socioeconomic disadvantage.

## Data Availability

All data produced in the present study are available upon reasonable request to the authors

## REFERENCES

1. Mathur R, Rentsch CT, Morton CE, Hulme WJ, Schultze A, MacKenna B, et al. Ethnic differences in SARS-CoV-2 infection and COVID-19-related hospitalisation, intensive care unit admission, and death in 17 million adults in England: an observational cohort study using the OpenSAFELY platform. Lancet. 2021 May 8;397(10286):1711–24.

2. Saatci D, Ranger TA, Garriga C, Clift AK, Zaccardi F, Tan PS, et al. Association Between Race and COVID-19 Outcomes Among 2.6 Million Children in England. JAMA Pediatr. 2021 Sep 1;175(9):928–38.

3. Lieberman-Cribbin W, Galanti M, Shaman J. Socioeconomic Disparities in Severe Acute Respiratory Syndrome Coronavirus 2 Serological Testing and Positivity in New York City. Open Forum Infect Dis. 2021 Dec;8(12):ofab534.

4. Metzger GA, Asti L, Quinn JP, Chisolm DJ, Xiang H, Deans KJ, et al. Association of the Affordable Care Act Medicaid Expansion with Trauma Outcomes and Access to Rehabilitation among Young Adults: Findings Overall, by Race and Ethnicity, and Community Income Level. J Am Coll Surg. 2021 Dec;233(6):776–93 e16.

5. K CM, Oral E, Straif-Bourgeois S, Rung AL, Peters ES. The effect of area deprivation on COVID-19 risk in Louisiana. Plos One. 2020;15(12):e0243028.

6. Hatef E, Chang HY, Kitchen C, Weiner JP, Kharrazi H. Assessing the Impact of Neighborhood Socioeconomic Characteristics on COVID-19 Prevalence Across Seven States in the United States. Front Public Health. 2020;8:571808.

7. Etowa J, Demeke J, Abrha G, Worku F, Ajiboye W, Beauchamp S, et al. Social determinants of the disproportionately higher rates of COVID-19 infection among African Caribbean and Black (ACB) population: A systematic review protocol. J Public Health Res. 2021 Dec 30;11(2).

8. Oluyomi AO, Gunter SM, Leining LM, Murray KO, Amos C. COVID-19 Community Incidence and Associated Neighborhood-Level Characteristics in Houston, Texas, USA. Int J Environ Res Public Health. 2021 Feb 4;18(4).

9. Dong Y, Mo X, Hu Y, Qi X, Jiang F, Jiang Z, et al. Epidemiology of COVID-19 Among Children in China. Pediatrics. 2020 Jun;145(6).

10. Rao S, Lee GM, Razzaghi H, Lorman V, Mejias A, Pajor NM, et al. Clinical Features and Burden of Postacute Sequelae of SARS-CoV-2 Infection in Children and Adolescents. JAMA Pediatr. 2022 Oct 1;176(10):1000–9.

11. Pfaff ER, Girvin AT, Bennett TD, Bhatia A, Brooks IM, Deer RR, et al. Identifying who has long COVID in the USA: a machine learning approach using N3C data. Lancet Digit Health. 2022 Jul;4(7):e532–e41.

12. Zhang Y, Hu H, Fokaidis V, Lewis VC, Xu J, Zang C, et al. Identifying Contextual and Spatial Risk Factors for Post-Acute Sequelae of SARS-CoV-2 Infection: An EHR-based Cohort Study from the RECOVER Program. medRxiv. 2022 Oct 13.

13. Pfaff ER, Madlock-Brown C, Baratta JM, Bhatia A, Davis H, Girvin A, et al. Coding Long COVID: Characterizing a new disease through an ICD-10 lens. medRxiv. 2022 Sep 2.

14. Hill E, Mehta H, Sharma S, Mane K, Xie C, Cathey E, et al. Risk Factors Associated with Post-Acute Sequelae of SARS-CoV-2 in an EHR Cohort: A National COVID Cohort Collaborative (N3C) Analysis as part of the NIH RECOVER program. medRxiv. 2022 Aug 17.

15. Mejias A, Schuchard J, Rao S, Bennett DT, Jhaveri R, Thacker D, et al. Leveraging serology testing to identify children at risk for post-acute sequelae of SARS-CoV-2 infection: An EHR-based cohort study from the RECOVER program. medRxiv. 2022.

16. Forrest CB, Margolis PA, Bailey LC, Marsolo K, Del Beccaro MA, Finkelstein JA, et al. PEDSnet: a National Pediatric Learning Health System. J Am Med Inform Assoc. 2014 Jul-Aug;21(4):602–6.

17. Forrest CB, Margolis P, Seid M, Colletti RB. PEDSnet: how a prototype pediatric learning health system is being expanded into a national network. Health Aff (Millwood). 2014 Jul;33(7):1171–7.

18. Forrest CB, Burrows EK, Mejias A, Razzaghi H, Christakis D, Jhaveri R, et al. Severity of Acute COVID-19 in Children <18 Years Old March 2020 to December 2021. Pediatrics. 2022 Apr 1;149(4).

19. Kind AJH, Buckingham WR. Making Neighborhood-Disadvantage Metrics Accessible - The Neighborhood Atlas. N Engl J Med. 2018 Jun 28;378(26):2456–8.

20. Fritz CQ, Fleegler EW, DeSouza H, Richardson T, Kaiser SV, Sills MR, et al. Child Opportunity Index and Changes in Pediatric Acute Care Utilization in the COVID-19 Pandemic. Pediatrics. 2022 May 1;149(5).

21. Simon TD, Cawthon ML, Stanford S, Popalisky J, Lyons D, Woodcox P, et al. Pediatric medical complexity algorithm: a new method to stratify children by medical complexity. Pediatrics. 2014 Jun;133(6):e1647–54.

22. Simon TD, Cawthon ML, Popalisky J, Mangione-Smith R, Center of Excellence on Quality of Care Measures for Children with Complex N. Development and Validation of the Pediatric Medical Complexity Algorithm (PMCA) Version 2.0. Hosp Pediatr. 2017 Jul;7(7):373–7.

23. Bacong AM, Haro-Ramos AY. Willingness to Receive the COVID-19 Vaccine in California: Disparities by Race and Citizenship Status. J Racial Ethn Health Disparities. 2022 Nov 30.

24. Gonzalez M, Zeidan J, Lai J, Yusuf A, Wright N, Steiman M, et al. Socio-demographic disparities in receipt of clinical health care services during the COVID-19 pandemic for Canadian children with disability. BMC Health Serv Res. 2022 Nov 28;22(1):1434.

25. Taghrir MH, Akbarialiabad H, Abdollahi A, Ghahramani N, Bastani B, Paydar S, et al. Inequity and disparities mar existing global research evidence on Long COVID. Glob Health Promot. 2022 Aug 12:17579759221113276.

26. Bellandi D. Estimate: 10.5 Million Children Lost a Parent, Caregiver to COVID-19. JAMA. 2022 Oct 18;328(15):1490.

27. Tung EL, Peek ME, Rivas MA, Yang JP, Volerman A. Association Of Neighborhood Disadvantage With Racial Disparities In COVID-19 Positivity In Chicago. Health Aff (Millwood). 2021 Nov;40(11):1784–91.

28. Dawood FS, Porucznik CA, Veguilla V, Stanford JB, Duque J, Rolfes MA, et al. Incidence Rates, Household Infection Risk, and Clinical Characteristics of SARS-CoV-2 Infection Among Children and Adults in Utah and New York City, New York. JAMA Pediatr. 2022 Jan 1;176(1):59–67.

